# Dental Distrust and Discrimination: A Nationally Representative Perspective on LGBTQ+ Adults’ Experiences with Oral Health Care

**DOI:** 10.64898/2026.05.01.26352238

**Authors:** Morgan Santoro, Lisa J. Heaton, Hannah J. Cheung, Rebecca A. Preston, Eric P. Tranby

## Abstract

**Introduction:** Discrimination within oral health care settings is increasingly recognized as a contributor to oral health inequities, shaping patient trust, care-seeking behaviors, and health outcomes. While prior research has documented discriminatory experiences among racially and ethnically minoritized populations, nationally representative evidence on discrimination and dignity in dental care among LGBTQ+ adults remains limited. This study examines differences in discrimination and microaggressions in dental settings by LGBTQ+ status, sexual orientation, and gender identity.

**Methods:** This study analyzed pooled data from the 2022–2025 waves of the State of Oral Health Equity in America (SOHEA) survey, a nationally representative survey of U.S. adults aged 18 and older. Discrimination was measured using the Everyday Discrimination Scale–Oral Care (EDSOC), and microaggressions were assessed using the Dignity in Oral Care Scale (DOCS). Descriptive and bivariate analyses compared mean scores across identity groups. Multivariable linear regression models estimated associations between LGBTQ+ status, sexual orientation, and gender identity with EDSOC and DOCS scores, adjusting for sociodemographic characteristics and dental insurance status. Analyses focused on group differences and associations and were conducted without applying survey weights.

**Results:** The analytic sample included 15,591 adults from the 2022–2025 SOHEA surveys with complete data (52.5% of the total N=29,679); 12% identified as LGBTQ+. Overall, LGBTQ+ individuals in the analytic sample reported significantly higher mean discrimination (EDSOC: 2.97, SD=4.99) and microaggression (DOCS: 2.19, SD=3.22) scores than non-LGBTQ+ individuals (EDSOC: 1.72, SD=3.79; DOCS: 1.62, SD=2.80; p<0.001). Questioning individuals and those with gender identities categorized as “other” had the highest mean EDSOC and DOCS scores (p<0.001). In adjusted models controlling for sociodemographic and insurance factors, LGBTQ+ identity remained significantly associated with higher EDSOC (β=0.16, 95% CI=0.11–0.21) and DOCS (β=0.08, 95% CI=0.03–0.13) scores. Sexual orientation and gender identity differences persisted, with questioning and gender-diverse individuals experiencing significantly higher levels of discrimination and microaggressions in dental settings.

**Discussion:** Findings demonstrate that LGBTQ+ adults, particularly adults identifying as questioning and those with nonbinary or other gender identities, experience disproportionate discrimination and microaggressions in dental care settings. Addressing interpersonal and structural sources of bias in oral health care is critical to advancing equity and improving access to respectful, high-quality care for LGBTQ+ populations.

## Introduction

Discrimination within oral health care settings is increasingly identified as a critical driver of oral health inequities, shaping not only patients’ clinical experiences but also their trust, care-seeking behaviors, and long-term health outcomes. Experiences of microaggressions, feelings of disregard, and differential treatment from providers during dental visits have been linked to poorer self-rated oral health, decreased dental utilization, and delayed or foregone dental care (1, 2). In a nationally representative survey of U.S. adults, Raskin et al. demonstrated that racism and everyday discrimination during prior dental visits were associated with worser self-rated oral health, longer intervals between dental care, and a reduced likelihood of planning future preventive visits (3). Population -level evidence further supports these findings as well. Using Behavioral Risk Factor Surveillance System data, Wael Sabbah and colleagues found that emotional impact of racial discrimination significantly predicted a lower likelihood of dental visits among U.S. adults (4). Together, these studies underscore that oral health inequities are not produced solely by structural barriers to access to oral health care but are also perpetuated through discriminatory interpersonal dynamics within care.

Emerging research also highlights mechanisms through which discrimination in oral health care contributes to reduced utilization. Sokoto et al. found that experiences of racism in dental settings were significant predictors of dental care–related fear and anxiety, which in turn predicted lower dental utilization among Black/African American women (2). The relationship between racism in oral health care settings and dental utilization was observed only when mediated by dental fear and anxiety, suggesting that discriminatory encounters may discourage care by fostering emotional distress and avoidance. Evidence of discrimination shaping oral health behaviors is not limited to the United States. In a Brazilian study, researchers found that there was a negative association with preventive dental attendance and discrimination based on social class (5). Additionally, researchers in Australia noted that perceived racial discrimination was associated with higher frequencies of poor oral health in Australian adults, concluding that socially marginalized groups have a higher burden of oral health effects based on racial discrimination (6). Collectively, these findings highlight discrimination as a pervasive and consequential determinant of oral health across diverse contexts.

While a growing body of research has documented oral health outcomes among racially minoritized populations, less is known about how discrimination, specifically in oral health care settings, can affect outcomes for sexual and gender minoritized adults. An analysis of nationally representative data from the Kaiser Family Foundation Survey of Racism, Discrimination, and Health observed that LGBTQ+ (lesbian, gay, bisexual, transgender, queer/questioning, and other identities (7)) adults experienced significantly higher rates of discrimination across healthcare settings compared to non-LGBTQ+ adults, which included reports of receiving poorer treatment, feelings of mistrust of providers, and avoiding dental care due to prior negative encounters (8). A study examining discrimination in medical settings found that LGBTQ+ patients had reported higher levels of perceived discrimination by their providers, lower overall patient satisfaction, and worse physical health outcomes in comparison to their heterosexual and cisgender counterparts (9). These studies suggest that LGBTQ+ populations encounter systemic barriers within healthcare environments that potentially undermine trust and lower engagement, factors that are important for preventive and routine care.

Less is known about how these interpersonal dynamics manifest specifically within oral health care. Empirical studies examining LGBTQ+ patient experiences in dental settings are scarce and geographically limited. One of the few available studies, conducted by Tharp and colleagues, documented discomfort, perceived unfair treatment, and barriers to care among LGBTQ+ patients in oral health settings in Indiana and Michigan (10). However, the researchers noted the lack of broader, population-based data to draw causal conclusions. Narrative reviews further underscore this gap, concluding that LGBTQ+ oral health disparities, particularly experiences of discrimination within dental care, remain largely underexplored in national surveillance and epidemiologic research (11). Recent descriptive reports from CareQuest Institute for Oral Health using data from the nationally representative State of Oral Health Equity in America survey have highlighted disparities and inequities among LGBTQ+ populations, including LGBTQ+ people of color, emphasizing the need for analyses of large-scale data sets to examine these disparities on a national level (12, 13).

Evidence from other healthcare contexts provides reason for concern that compounded inequities may exist in oral health care. In 2020, The National Academies of Sciences, Engineering, and Medicine documented how structural and interpersonal stigma, including misgendering, exclusionary policies, and lack of provider cultural competence, erode trust and reduced healthcare utilization among LGBTQ+ populations (14). Given that dental care is considered invasive and dependent on patient–provider trust, the lack of robust data on LGBTQ+ experiences in dental settings represents a critical gap in oral health equity research. Addressing this gap is essential for understanding how discrimination and distrust may shape oral health behaviors and outcomes among LGBTQ+ adults and for informing inclusive, patient-centered approaches to dental care delivery.

The objective of this study was to examine discrimination and microaggressions experienced in the dental setting by individuals identifying as part of the LGBTQ+ community. Bivariate analyses of the nationally representative data from the State of Oral Health Equity in America (SOHEA) survey were used to examine differences discrimination and microaggression experiences by LGBTQ+ status, sexual orientation type, and gender identity, and multivariable models examined these differences while adjusting for key demographic variables. It was hypothesized that individuals identifying as part of the LGBTQ+ community, as sexual orientation minorities, or as gender identity minorities would report more discrimination and microaggressions than non-LGBTQ+ individuals.

## Materials and Methods

### Study Design and Data Source

Data for this study came from the 2022-2025 waves of the annual State of Oral Health Equity in America (SOHEA) survey. SOHEA is a nationally representative survey designed to assess adults’ oral health–related attitudes, experiences, and behaviors (1, 3, 15). Collaborators from CareQuest Institute for Oral Health developed the survey with feedback from external collaborators and was pre-tested (48 adults in December 2021 for the 2022 round; 119 adults in January 2023 for the 2023 round; 25 in February 2024 for the 2024 round, and 59 in December 2024 for the 2025 round). The survey was administered online (96.7% in 2022; 96.2% in 2023; 96.2% in 2024; 97.3% in 2025) and via telephone (3.3% in 2022, 3.8% in 2023; 3.8% in 2024; 2.7% in 2025) in four rounds in January–February 2022 and 2025, February–March 2023, and March–May 2024, with adults aged 18 and older by NORC at the University of Chicago as part of the AmeriSpeak panel (16, 17). This study protocol was reviewed and determined to be exempt from Human Subjects review by the WCG Institutional Review Board.

AmeriSpeak is a probability-based panel designed to be representative of the U.S. household population (17). Sampled households were recruited via U.S. mail, telephone, and in-person field interviews. In the 2022 survey round, a sampling frame of 17,603 households yielded a final analytic sample of 5,682 respondents, corresponding to a survey completion rate of 32.3%. In the 2023 round, the sampling frame, final sample, and completion rate were 18,521 households, 5,240 respondents, and a 28.3% completion rate. In 2024, these figures corresponded to 22,448 households, 9,307 respondents, and a 41.5% response rate; in 2025, there were 19,193 households, 9,450 respondents, and a 49.2% response rate.

### Participants

Participants were adults aged 18 and above participating in the SOHEA survey through the Amerispeak panel. Each adult responded on behalf of themselves and others in their household. Adults were ineligible to participate if they were under the age of 18 or if someone living in their household had already participated in that year’s SOHEA survey.

### Dependent Variables

Respondents reported their experiences with discrimination in the dental setting using the Everyday Discrimination Scale—Oral Care (EDSOC). The EDSOC is a 7-item scale that was adapted from a medical version ((18) of the Everyday Discrimination Scale (19). The EDSOC was adapted from the medical scale for relevance to the dental setting (20). The 7 EDSOC items ask respondents to indicate how frequently they experienced a discriminatory situation in an oral health care setting within the prior year (e.g., “How often have any of the following things happened to you in the last year? You felt that a dental provider or a member of the dental team acted as if they were better than you”) on a 5-point scale (0 = never; 1 = rarely; 2 = sometimes; 3 = mostly; 4 = always). Total scores range from 0 to 28, with higher scores indicating more discriminatory experiences in the dental setting. The EDSOC has been shown to have good internal reliability (Cronbach’s alpha = 0.88)(20). Mean EDSOC scores were calculated for groups based on sexual orientation and gender identity.

Participants reported their experiences with microaggressions (i.e., being treated without dignity) in the dental setting using the Dignity in Oral Care Scale (DOCS). The DOCS is a 4-item scale that asks respondents how much they agree with statements regarding their most recent dental visit (1 = strongly agree; 2 = somewhat agree; 3 = neither agree nor disagree; 4 = somewhat disagree; 5 = strongly disagree). For example, respondents are asked, “At my last oral health visit, my dental provider respected me.” The DOCS is scored such that higher item scores represent stronger disagreement with positively worded items; total scores range from 4 to 20, with higher scores representing more experiences with microaggressions and less dignity. The DOCS has been shown to have excellent internal reliability (Cronbach’s alpha = 0.92) (20).Mean DOCS scores were calculated for groups based on sexual orientation and gender identity.

### Independent Variables and Covariates

Respondents reported their sexual orientation as bisexual, gay/lesbian, questioning, other, or straight, and indicated their gender identity as female, male, transgender, or other. Throughout this article, the abbreviation “LGBTQ+” is used to specify the sexual orientations and gender identities that were measured specifically (i.e., lesbian, gay, bisexual, transgender, questioning/queer, and other (+)) (7). Covariates included age in years (18-29, 30-44, 45-59, or 60 or above), race/ethnicity (Asian/Pacific Islander, non-Hispanic, Black/African American, non-Hispanic, Hispanic/Latino/a/x, Other, non-Hispanic, or White, non-Hispanic), annual household income (less than $30,000, $30,000 to under $60,000, $60,000 to under $100,000, or $100,000 or more), highest education level achieved (less than high school, high school graduate or equivalent, vocational/associate’s degree, bachelor’s degree, or postgraduate/professional degree), employment status (working – paid employee or self-employed or not working – retired, disabled, or other), and dental insurance status (have dental insurance or do not have dental insurance).

### Statistical Analyses

Descriptive analyses were conducted to analyze and summarize the overall sample variables. Means and standard deviations were calculated for continuous variables, including the EDSOC and DOCS scales. Categorical variables, such as gender identity, sexual orientation, LGBTQ+ status, age category, race/ethnicity, income, education, dental insurance coverage, and employment status, were summarized using counts and percentages. Due to the analytical goal of examining group differences rather than producing population-level estimates, all statistics were generated without applying survey weights. Unweighted analyses were selected to improve statistical precision and stability, particularly given the focus on LGBTQ+ status, gender identity and sexual orientation. Mean EDSOC and DOCS scores (with standard deviations) were calculated to compare between LGBTQ+ and non-LGBTQ+ groups. Data were excluded for respondents who did not have complete EDSOC or DOCS responses or did not respond to either gender identity or sexual orientation questions.

To examine differences in discriminatory treatment and dignity experiences across LGBTQ+ status, sexual orientation groups, and gender identity, a series of unweighted descriptive analyses were conducted. Continuous variables, including the EDSOC and DOCS scales, were compared between two groups using the Wilcoxon rank-sum test or across multiple groups using the Kruskal-Wallis rank-sum test, given the non-normal distribution of these measures. Categorical variables, such as age category, race/ethnicity, income, education, employment, and dental insurance status, were compared across groups using Person’s chi-square test. All tests assessed overall group differences without adjustment for multiple comparisons, and a two-sided p-value threshold of 0.05 was used to evaluate statistical significance.

A series of multivariable linear regression models were estimated to examine the association between the primary identity variables with EDSOC and DOCS scores. Each model included one primary identity variable of interest (e.g., sexual orientation identity, gender identity, or LGBTQ+ status) while adjusting for sociodemographic characteristics, including age category, race/ethnicity, household income, education level, dental insurance status, and employment status. Models were fit using unweighted data, consistent with the analytical goal of identifying group differences rather than generating population-level estimates. Results were reported as β (beta) coefficients, 95% confidence intervals, p-values, and r-squared.

## Results

The study sample consisted of 15,591 adults who participated in the 2022-2025 rounds of the SOHEA survey who had complete data (52.5% of the entire sample of N=29,679 from across four years); Table 1 presents the demographic characteristics of the study sample. Twelve percent of the sample identified as LGBTQ+. In terms of sexual orientation, the largest percentage represented after heterosexual/straight (88%) was bisexual (4.4%). More than half of respondents (56%) identified their gender identity as female, 43% as male, 0.6% as other, and 0.2% as transgender. The highest percentage of respondents with complete data were aged 60 or above (42%), white, non-Hispanic (70%), had a vocational/technical/associate’s degree or some college (37%), were employed (56%), and had dental insurance (72%; Table 1). Annual household income was equally distributed across four categories.

**Table 1.**
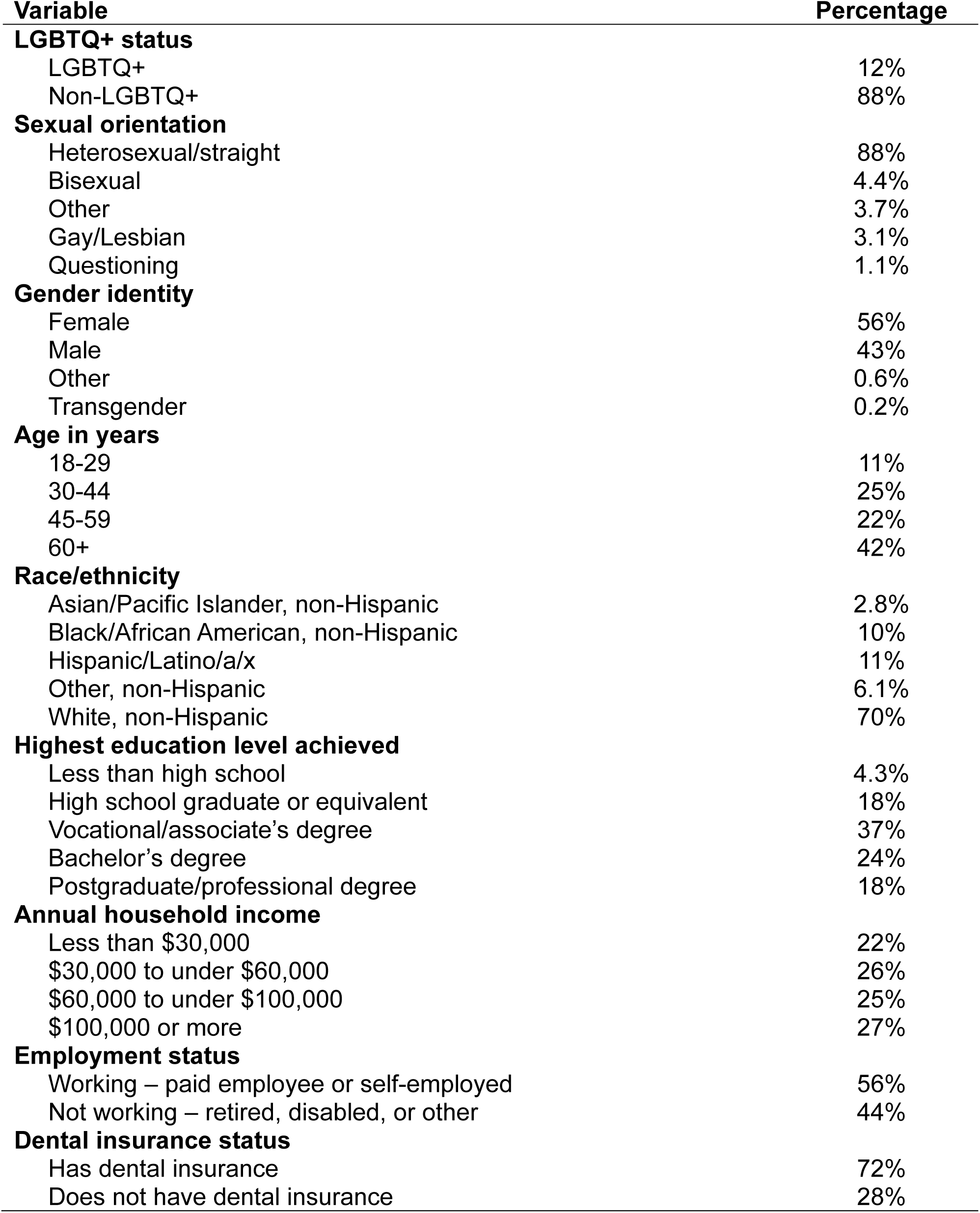
Demographic Characteristics of the Study Sample (N=15,591)

Figure 1 shows EDSOC (discrimination) and DOCS (microaggression) scores by LGBTQ+ status, sexual orientation, and gender identity. Individuals identifying as LGBTQ+ had statistically significantly higher EDSOC (mean = 2.97, standard deviation (sd) = 4.99)) and DOCS (mean = 2.19, sd = 3.22) scores than non-LGBTQIA individuals (EDSOC mean = 1.72 (sd = 3.79), DOCS mean = 1.62 (sd = 2.80, p<0.001). Regarding sexual orientation, individuals identifying as questioning had significantly higher mean EDSOC (mean = 4.92 (sd = 6.57)) and DOCS (mean = 2.90 (sd = 3.44)) scores than individuals identifying as heterosexual/straight (EDSOC mean = 1.70 (sd = 3.76), DOCS mean = 1.61 (sd = 2.79, p<0.001)). In terms of gender identity, individuals identifying as other had significantly higher EDSOC (mean = 5.75 (sd = 6.32)) and DOCS (mean = 3.78 (sd = 4.10)) scores than individuals identifying as female (EDSOC mean = 1.88 (sd = 3.92), DOCS mean = 1.61 (sd = 2.82, p<0.001)) and significantly higher DOCS (mean = 3.78 (sd = 4.10)) scores than individuals identifying as male (mean = 1.74 (sd = 2.85, p<0.001)).

**Figure 1:**
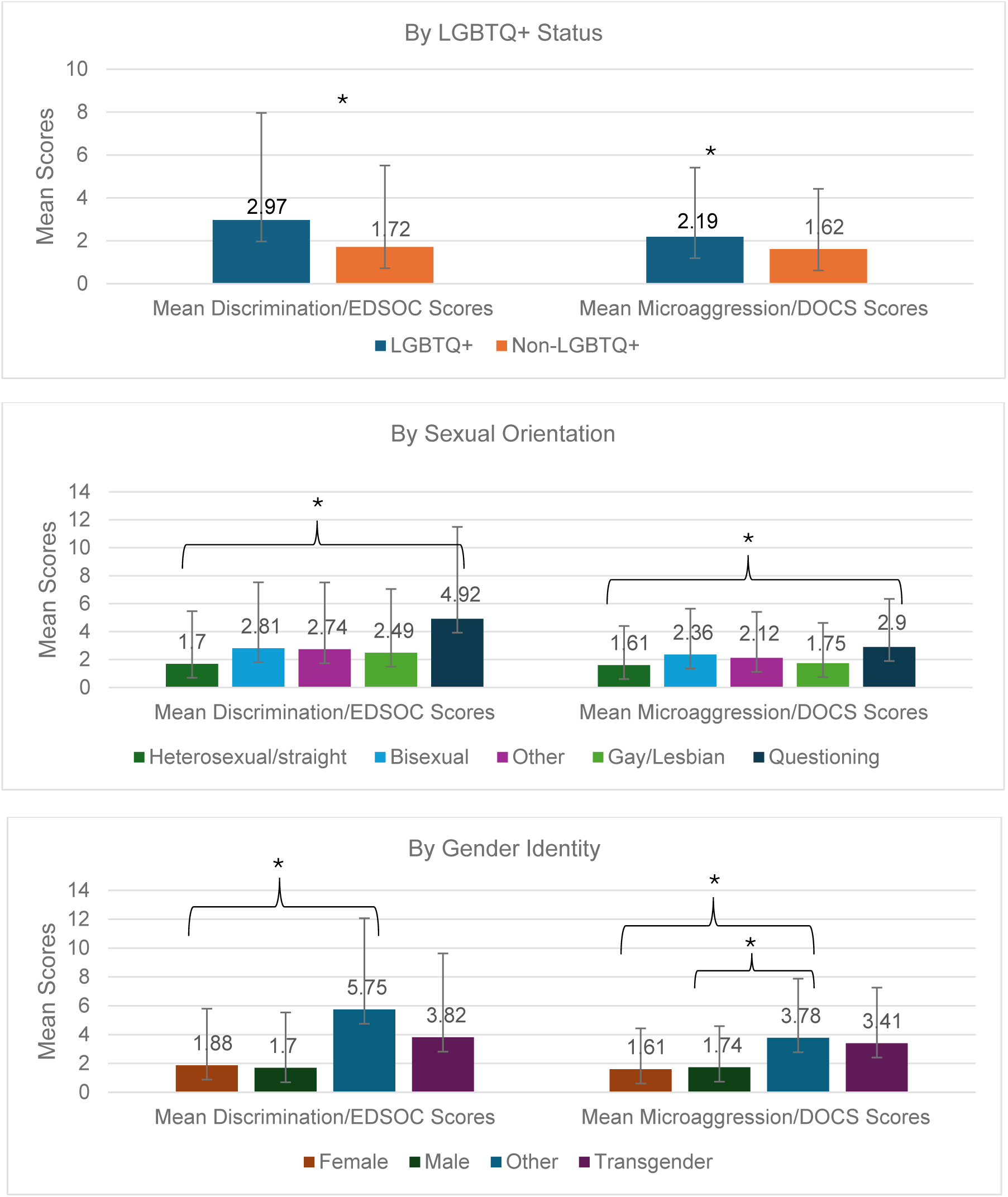
Mean EDSOC and DOCS Scores by LGBTQ+ Status, Sexual Orientation and Gender Identity

After controlling for the confounding variables of age in years, race/ethnicity, highest education level achieved, annual household income, employment status, and dental insurance status, individuals identifying as LGBTQ+ scored significantly higher on the EDSOC measure than non-LGBTQ+ individuals (β = 0.16, 95% confidence interval (CI) = 0.11, 0.21; p<0.001; Table 2; Appendices A-C). Compared to individuals identifying as heterosexual/straight, those identifying their sexual orientation as other, gay/lesbian, or questioning scored significantly higher on the EDSOC measure, indicating experiencing more discriminatory treatment in dental settings (p’s < 0.01). Individuals identifying their gender identity as other scored significantly higher on the EDSOC measure than individuals identifying as male (β = 0.64, 95% CI = 0.45, 0.84; p<0.001).

**Table 2:** Multivariable Regression Models for Discrimination/EDSOC Scores.

| **Variable** | **Beta** | **95% CI** | **p-value** | **R<sup>2</sup>** |
| --- | --- | --- | --- | --- |
| **LGBTQ+ Status** |  |  |  | 0.12 |
| Non-LGBTQ+ | ref | ref | ref |  |
| *LGBTQ+* | *0.16* | *0.11, 0.21* | *<0.001* |  |
| **Sexual orientation** |  |  |  | 0.13 |
| Heterosexual/straight | ref | ref | ref |  |
| Bisexual | 0.04 | -0.04, 0.11 | 0.3 |  |
| *Gay/Lesbian* | *0.13* | *0.05, 0.22* | *0.002* |  |
| *Other* | *0.15* | *0.07, 0.23* | *<0.001* |  |
| *Questioning* | *0.62* | *0.48, 0.77* | *<0.001* |  |
| **Gender identity** |  |  |  | 0.12 |
| Male | ref | ref | ref |  |
| Female | -0.02 | -0.05, 0.01 | 0.2 |  |
| *Other* | *0.64* | *0.45, 0.84* | *<0.001* |  |
| Transgender | 0.24 | -0.07, 0.55 | 0.13 |  |

After controlling for confounding variables, individuals identifying as LGBTQ+ scored significantly higher on the DOCS measure than non-LGBTQ+ individuals (β = 0.08, 95% CI = 0.03, 0.13; p = 0.003; Table 3; Appendices D-F). Compared to individuals identifying as heterosexual/straight, those identifying as questioning scored significantly higher on the DOCS measure (β = 0.29, 95% CI = 0.13, 0.46; p<0.001), indicating experiencing more microaggressions in dental settings. Individuals identifying their gender identity as female scored significantly lower on the DOCS measure (β = -0.12, 95% CI = -0.15, -0.88; p<0.001) than those identifying as male. Meanwhile, individuals identifying their gender identity as other had significantly higher DOCS scores than males (β = 0.42, 95% CI = 0.18, 0.66, p<0.001).

**Table 3:**
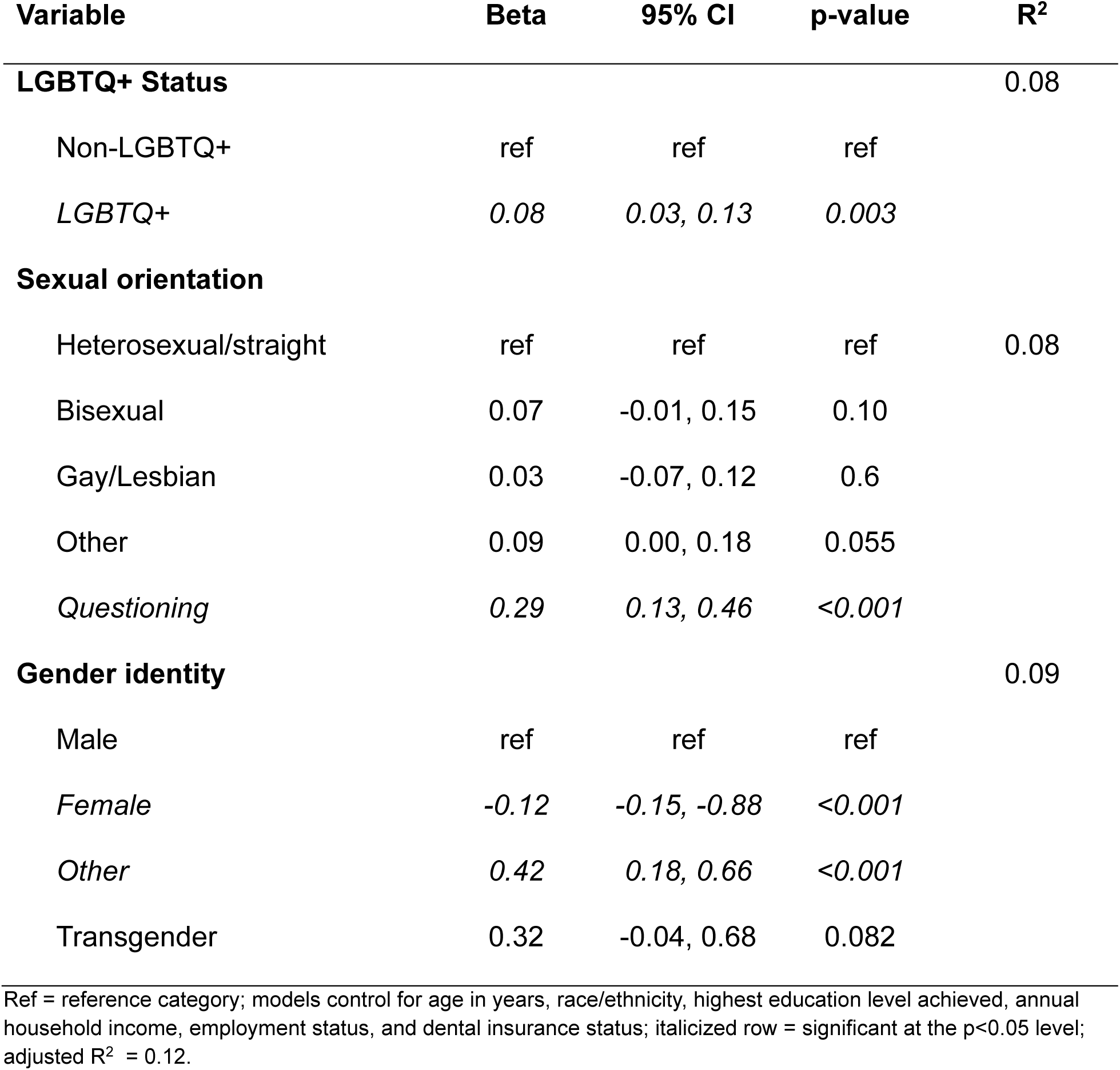
Multivariable Regression Models for Microaggression/DOCS Scores.

| Variable | Beta | 95% CI | p-value | R<sup>2</sup> |
| --- | --- | --- | --- | --- |
| **LGBTQ+ Status** |  |  |  | 0.08 |
| Non-LGBTQ+ | ref | ref | ref |  |
| *LGBTQ+* | *0.08* | *0.03, 0.13* | *0.003* |  |
| **Sexual orientation** |  |  |  |  |
| Heterosexual/straight | ref | ref | ref | 0.08 |
| Bisexual | 0.07 | -0.01, 0.15 | 0.10 |  |
| Gay/Lesbian | 0.03 | -0.07, 0.12 | 0.6 |  |
| Other | 0.09 | 0.00, 0.18 | 0.055 |  |
| *Questioning* | *0.29* | *0.13, 0.46* | *<0.001* |  |
| **Gender identity** |  |  |  | 0.09 |
| Male | ref | ref | ref |  |
| *Female* | *-0.12* | *-0.15, -0.88* | *<0.001* |  |
| *Other* | *0.42* | *0.18, 0.66* | *<0.001* |  |
| Transgender | 0.32 | -0.04, 0.68 | 0.082 |  |

Ref = reference category; models control for age in years, race/ethnicity, highest education level achieved, annual household income, employment status, and dental insurance status; italicized row = significant at the p<0.05 level; adjusted R<sup>2</sup> = 0.12.

## Discussion

After adjusting for sociodemographic and insurance-related factors, individuals identifying as LGBTQ+ reported significantly higher levels of both discriminatory treatment and microaggressions in dental settings compared with non-LGBTQ+ individuals. Sexual orientation and gender identity differences were evident, with gay/lesbian, questioning, and gender-diverse individuals experiencing higher discrimination and microaggressions, while individuals identifying as female reported fewer microaggressions than males. Overall, the findings indicate persistent disparities in dental care experiences associated with LGBTQ+ identity.

The findings of this study support and expand prior work that shows substantial impact of discrimination and microaggressions in the dental setting on individuals from racial/ethnic minoritized groups (1, 2, 4, 6). A scoping review demonstrated implicit bias among oral health care providers, faculty, and students toward individuals from racial/ethnic minoritized groups that may result in poorer oral health treatment outcomes (21).Given known evidence of stigma in oral health care toward minoritized groups, it is critical to understand the prevalence and impact of such discrimination faced by individuals identifying as LGBTQ+.

When examining discriminatory treatment and microaggression experiences across LGBTQ+ status, sexual orientation groups, and gender identity, a multi-year, large nationally representative sample, and consistent survey design over multiple years with previously validated dependent measures yielded comparison and significant results. Additionally, previous research demonstrated the need for detailed sexual orientation and gender identity categories to reduce misclassification and increase representation (22, 23). SOHEA’s survey design collects both binary (male/female) and non-binary (transgender, other) gender identity, categories sexual orientation as bisexual, gay/lesbian, questioning, other, or straight, and uses gender neutral terminology in survey questions. Further research and evaluation should focus on the adoption of standardized responses for gender identity and sexual orientation.

Some limitations should be considered when interpreting these results. The cross-sectional design of the survey design does not allow for causal conclusions to be drawn. Additionally, the data collected in this survey are self-reported; responses may be affected by recall bias and reporting bias possibly due to societal changes, current health challenges, and social desirability responsiveness. While the survey is nationally representative, analyses were conducted without applying survey weights; therefore, findings should be interpreted as analytic associations and may not be fully generalizable to the U.S. LGBTQ+ population. Furthermore, small, absolute numbers in transgender and other identity categories limit comparability and may have joined identities and experiences into one category (24, 25).

The findings of this study emphasize the critical need for inclusive, affirming dental practice environments in which patients of all sexual orientations and gender identities feel comfortable and accepted. As experiencing discrimination in the dental setting is linked to poor self-rated oral health and avoidance of dental care (20), providing oral health care that is free of discriminatory treatment and microaggressions is key to ensuring that LGBTQ+ individuals are able to receive care that optimizes their oral health.

Providing such culturally responsive care begins with oral health education programs; a recent climate survey by the American Dental Education Association (ADEA) found that dental students, allied dental profession students, staff, faculty, and administrators identifying as LGBTQ+ reported less satisfaction with their academic program and employment and more experiences with discrimination and harassment than their non-LGBTQIA counterparts (26, 27). While many dental schools signal broad support for diversity, LGBTQ+ inclusion, especially in curriculum and clinical training, remains limited, inconsistent, and largely informal, leaving graduates underprepared to meet the oral health needs of queer and transgender patients (28). Increasing representation of LGBTQ+ staff, faculty, and administration in oral health education programs is key to improving a sense of inclusion and belonging within the educational setting and beyond (29). Pre-doctoral, post-doctoral, and continuing education programs aimed at increasing cultural responsiveness in providing care to LGBTQ+ patients should be included as part of training and licensure requirements (11).

## Conclusions

LGBTQ+ adults, especially questioning and other identity groups, experience disproportionate discrimination and microaggressions in dental care compared to non-LGBTQ+ adults. As experiencing discrimination and microaggressions in dental care is linked with poor oral health outcomes, it is critical to address provider biases and structural inequities to provide culturally responsive, supportive, and inclusive care to LGBTQ+ individuals.

## Data Availability

All data produced in the present study are available upon reasonable request to the authors.

---

**Appendix B:** Multivariable Regression Models for Discrimination/EDSOC Scores by LGBTQ+ Status.

Appendix A: Multivariable Regression Models for Discrimination/EDSOC Scores by LGBTQ+ Status
| Variable | Beta | 95% CI | p-value |
| --- | --- | --- | --- |
| **LGBTQ+ Status** |  |  |  |
| Non-LGBTQ+ | ref | ref | ref |
| *LGBTQ+* | *0.16* | *0.11, 0.21* | *<0.001* |
| **Age in years** |  |  |  |
| 18-29 | ref | ref | ref |
| 30-44 | 0.00 | -0.06, 0.05 | >0.9 |
| 45-59 | -0.20 | -0.26, -0.15 | <0.001 |
| 60+ | -0.41 | -0.46, -0.35 | <0.001 |
| **Race/ethnicity** |  |  |  |
| White | ref | ref | ref |
| *Asian* | *0.35* | *0.26, 0.44* | *<0.001* |
| *Black* | *0.22* | *0.17, 0.27* | *<0.001* |
| *Hispanic* | *0.28* | *0.23, 0.33* | *<0.001* |
| *Other* | *0.26* | *0.16, 0.32* | *<0.001* |
| **Annual household income** |  |  |  |
| Less than \$30,000 | ref | ref | ref |
| *\$30,000 to under \$60,000* | *-0.24* | *-0.29, -0.20* | *<0.001* |
| *\$60,000 to under \$100,000* | *-0.34* | *-0.38, -0.29* | *<0.001* |
| *\$100,000 or more* | *-0.39* | *-0.43, -0.34* | *<0.001* |
| **Highest education level achieved** |  |  |  |
| Less than high school | ref | ref | ref |
| *High school graduate/equivalent* | *-0.22* | *-0.30, -0.13* | *<0.001* |
| *Some college/associate's degree* | *-0.33* | *-0.41, -0.25* | *<0.001* |
| *Bachelor's degree* | *-0.41* | *-0.49, -0.32* | *<0.001* |
| *Postgraduate/professional degree* | *-0.37* | *-0.46, -0.29* | *<0.001* |
| **Employment** |  |  |  |
| Working – paid or self-employed | ref | ref | ref |
| Not working – retired/disabled/other | 0.00 | -0.03, 0.04 | 0.8 |
| **Dental insurance** |  |  |  |
| Have dental insurance | ref | ref | ref |
| *Do not have dental insurance* | *0.16* | *0.13, 0.20* | *<0.001* |

**Appendix B:** Multivariable Regression Models for Discrimination/EDSOC Scores by Sexual Orientation.

| Variable | Beta | 95% CI | p-value |
| --- | --- | --- | --- |
| **Sexual orientation** |  |  |  |
| Straight | ref | ref | ref |
| Bisexual | 0.04 | -0.04, .011 | 0.3 |
| *Gay/lesbian* | *0.13* | *0.05, 0.22* | *0.002* |
| *Other* | *0.15* | *0.07, 0.23* | *<0.001* |
| *Questioning* | *0.62* | *0.48, 0.77* | *<0.001* |
| **Age in years** |  |  |  |
| 18-29 | ref | ref | ref |
| 30-44 | -0.01 | -0.06, 0.05 | 0.8 |
| 45-59 | -0.21 | -0.27, -0.16 | <0.001 |
| 60+ | -0.41 | -0.47, -0.35 | <0.001 |
| **Race/ethnicity** |  |  |  |
| White | ref | ref | ref |
| *Asian* | *0.33* | *0.24, 0.43* | *<0.001* |
| *Black* | *0.22* | *0.17, 0.27* | *<0.001* |
| *Hispanic* | *0.27* | *0.22, 0.32* | *<0.001* |
| *Other* | *0.24* | *0.17, 0.30* | *<0.001* |
| **Annual household income** |  |  |  |
| Less than \$30,000 | ref | ref | ref |
| *\$30,000 to under \$60,000* | *-0.24* | *-0.29, -0.20* | *<0.001* |
| *\$60,000 to under \$100,000* | *-0.33* | *-0.38, -0.29* | *<0.001* |
| *\$100,000 or more* | *-0.38* | *-0.43, -0.33* | *<0.001* |
| **Highest education level achieved** |  |  |  |
| Less than high school | ref | ref | ref |
| *High school graduate/equivalent* | *-0.22* | *-0.30, -0.14* | *<0.001* |
| *Some college/associate's degree* | *-0.34* | *-0.42, -0.26* | *<0.001* |
| *Bachelor's degree* | *-0.42* | *-0.50, -0.33* | *<0.001* |
| *Postgraduate/professional degree* | *-0.38* | *-0.47, -0.30* | *<0.001* |
| **Employment** |  |  |  |
| Working – paid or self-employed | ref | ref | ref |
| Not working – retired/disabled/other | 0.00 | -0.04, 0.03 | >0.9 |
| **Dental insurance** |  |  |  |
| Have dental insurance | ref | ref | ref |
| *Do not have dental insurance* | *0.16* | *0.13, 0.20* | *<0.001* |

**Appendix C:** Multivariable Regression Models for Discrimination/EDSOC Scores by Gender Identity.

| Variable | Beta | 95% CI | p-value |
| --- | --- | --- | --- |
| **Gender identity** |  |  |  |
| Male | ref | ref | ref |
| Female | -0.02 | -0.05, 0.01 | 0.2 |
| *Other* | *0.64* | *0.45, 0.84* | *<0.001* |
| Transgender | 0.24 | -0.07, 0.55 | 0.13 |
| **Age in years** |  |  |  |
| 18-29 | ref | ref | ref |
| 30-44 | -0.02 | -0.08, 0.03 | 0.4 |
| 45-59 | -0.23 | -0.28, -0.17 | *<0.001* |
| 60+ | -0.42 | -0.48, -0.37 | *<0.001* |
| **Race/ethnicity** |  |  |  |
| White | ref | ref | ref |
| *Asian* | *0.33* | *0.24, 0.43* | *<0.001* |
| *Black* | *0.22* | *0.17, 0.27* | *<0.001* |
| *Hispanic* | *0.27* | *0.22, 0.32* | *<0.001* |
| *Other* | *0.25* | *0.19, 0.31* | *<0.001* |
| **Annual household income** |  |  |  |
| Less than \$30,000 | ref | ref | ref |
| *\$30,000 to under \$60,000* | *-0.23* | *-0.27, -0.18* | *<0.001* |
| *\$60,000 to under \$100,000* | *-0.33* | *-0.38, -0.28* | *<0.001* |
| *\$100,000 or more* | *-0.39* | *-0.44, -0.34* | *<0.001* |
| **Highest education level achieved** |  |  |  |
| Less than high school | ref | ref | ref |
| *High school graduate/equivalent* | *-0.21* | *-0.29, -0.13* | *<0.001* |
| *Some college/associate's degree* | *-0.32* | *-0.40, -0.24* | *<0.001* |
| *Bachelor's degree* | *-0.40* | *-0.48, -0.31* | *<0.001* |
| *Postgraduate/professional degree* | *-0.36* | *-0.45, -0.28* | *<0.001* |
| **Employment** |  |  |  |
| Working – paid or self-employed | ref | ref | ref |
| Not working – retired/disabled/other | 0.00 | -0.03, 0.04 | 0.8 |
| **Dental insurance** |  |  |  |
| Have dental insurance | ref | ref | ref |
| *Do not have dental insurance* | *0.16* | *0.12, 0.19* | *<0.001* |

Ref = reference category; models control for age in years, race/ethnicity, highest education level achieved, annual household income, employment status, and dental insurance status; italicized row = significant at the $p < 0.05$ level; adjusted $R^2 = 0.12$ .

**Appendix D:** Multivariable Regression Models for Microaggression/DOCS Scores by LGBTQ+ Status.

| Variable | Beta | 95% CI | p-value |
| --- | --- | --- | --- |
| **LGBTQ+ Status** |  |  |  |
| Non-LGBTQ+ | ref | ref | ref |
| *LGBTQ+* | *0.08* | *0.03, 0.13* | *0.003* |
| **Age in years** |  |  |  |
| 18-29 | ref | ref | ref |
| 30-44 | 0.01 | -0.06, 0.07 | 0.9 |
| 45-59 | -0.12 | -0.19, -0.06 | <0.001 |
| 60+ | -0.32 | -0.38, -0.26 | <0.001 |
| **Race/ethnicity** |  |  |  |
| White | ref | ref | ref |
| *Asian* | *0.23* | *0.13, 0.33* | *<0.001* |
| *Black* | *0.16* | *0.11, 0.22* | *<0.001* |
| *Hispanic* | *0.15* | *0.09, 0.20* | *<0.001* |
| *Other* | *0.16* | *0.09, 0.23* | *<0.001* |
| **Annual household income** |  |  |  |
| Less than \$30,000 | ref | ref | ref |
| *\$30,000 to under \$60,000* | *-0.15* | *-0.20, -0.10* | *<0.001* |
| *\$60,000 to under \$100,000* | *-0.27* | *-0.32, -0.22* | *<0.001* |
| *\$100,000 or more* | *-0.33* | *-0.39, -0.28* | *<0.001* |
| **Highest education level achieved** |  |  |  |
| Less than high school | ref | ref | ref |
| *High school graduate/equivalent* | *-0.20* | *-0.29, -0.10* | *<0.001* |
| *Some college/associate's degree* | *-0.29* | *-0.38, -0.20* | *<0.001* |
| *Bachelor's degree* | *-0.39* | *-0.49, -0.30* | *<0.001* |
| *Postgraduate/professional degree* | *-0.44* | *-0.54, -0.34* | *<0.001* |
| **Employment** |  |  |  |
| Working – paid or self-employed | ref | ref | ref |
| Not working – retired/disabled/other | 0.03 | -0.01, -0.07 | 0.11 |
| **Dental insurance** |  |  |  |
| Have dental insurance | ref | ref | ref |
| *Do not have dental insurance* | *0.17* | *0.13, 0.21* | *<0.001* |

Ref = reference category; models control for age in years, race/ethnicity, highest education level achieved, annual household income, employment status, and dental insurance status; italicized row = significant at the $p < 0.05$ level; adjusted $R^2 = 0.08$ .

**Appendix E:** Multivariable Regression Models for Microaggression/DOCS Scores by Sexual Orientation.

| Variable | Beta | 95% CI | p-value |
| --- | --- | --- | --- |
| **Sexual orientation** |  |  |  |
| Straight | ref | ref | ref |
| Bisexual | 0.07 | -0.01, 0.15 | 0.10 |
| Gay/lesbian | 0.03 | -0.07, 0.12 | 0.6 |
| Other | 0.09 | 0.00, 0.18 | 0.055 |
| *Questioning* | *0.29* | *0.13, 0.46* | *<0.001* |
| **Age in years** |  |  |  |
| 18-29 | ref | ref | ref |
| 30-44 | 0.00 | -0.06, 0.07 | >0.9 |
| 45-59 | -0.13 | -0.19, -0.06 | <0.001 |
| 60+ | -0.32 | -0.39, -0.26 | <0.001 |
| **Race/ethnicity** |  |  |  |
| White | ref | ref | ref |
| *Asian* | *0.23* | *0.13, 0.33* | *<0.001* |
| *Black* | *0.16* | *0.10, 0.21* | *<0.001* |
| *Hispanic* | *0.15* | *0.09, 0.20* | *<0.001* |
| *Other* | *0.16* | *0.09, 0.23* | *<0.001* |
| **Annual household income** |  |  |  |
| Less than \$30,000 | ref | ref | ref |
| *\$30,000 to under \$60,000* | *-0.15* | *-0.20, -0.10* | *<0.001* |
| *\$60,000 to under \$100,000* | *-0.27* | *-0.32, -0.21* | *<0.001* |
| *\$100,000 or more* | *-0.33* | *-0.39, -0.28* | *<0.001* |
| **Highest education level achieved** |  |  |  |
| Less than high school | ref | ref | ref |
| *High school graduate/equivalent* | *-0.21* | *-0.30, -0.11* | *<0.001* |
| *Some college/associate's degree* | *-0.30* | *-0.39, -0.21* | *<0.001* |
| *Bachelor's degree* | *-0.40* | *-0.50, -0.31* | *<0.001* |
| *Postgraduate/professional degree* | *-0.46* | *-0.55, -0.36* | *<0.001* |
| **Employment** |  |  |  |
| Working – paid or self-employed | ref | ref | ref |
| Not working – retired/disabled/other | 0.03 | -0.01, 0.07 | 0.12 |
| **Dental insurance** |  |  |  |
| Have dental insurance | ref | ref | ref |
| *Do not have dental insurance* | *0.16* | *0.12, 0.20* | *<0.001* |

Ref = reference category; models control for age in years, race/ethnicity, highest education level achieved, annual household income, employment status, and dental insurance status; italicized row = significant at the $p < 0.05$ level; adjusted $R^2 = 0.08$ .

**Appendix F:** Multivariable Regression Models for Microaggression/DOCS Scores by Gender Identity.

| Variable | Beta | 95% CI | p-value |
| --- | --- | --- | --- |
| **Gender identity** |  |  |  |
| Male | ref | ref | ref |
| *Female* | -0.12 | -0.15, -0.08 | <0.001 |
| *Other* | 0.42 | 0.18, 0.66 | <0.001 |
| Transgender | 0.32 | -0.04, -0.68 | 0.082 |
| **Age in years** |  |  |  |
| 18-29 | ref | ref | ref |
| 30-44 | 0.00 | -0.06, 0.06 | >0.9 |
| 45-59 | -0.13 | -0.19, -0.06 | <0.001 |
| 60+ | -0.33 | -0.39, -0.27 | <0.001 |
| **Race/ethnicity** |  |  |  |
| White | ref | ref | ref |
| *Asian* | 0.21 | 0.11, 0.31 | <0.001 |
| *Black* | 0.17 | 0.11, 0.23 | <0.001 |
| *Hispanic* | 0.14 | 0.09, 0.20 | <0.001 |
| *Other* | 0.16 | 0.09, 0.23 | <0.001 |
| **Annual household income** |  |  |  |
| Less than \$30,000 | ref | ref | ref |
| *\$30,000 to under \$60,000* | -0.15 | -0.20, -0.10 | <0.001 |
| *\$60,000 to under \$100,000* | -0.28 | -0.33, -0.23 | <0.001 |
| *\$100,000 or more* | -0.35 | -0.41, -0.30 | <0.001 |
| **Highest education level achieved** |  |  |  |
| Less than high school | ref | ref | ref |
| *High school graduate/equivalent* | -0.20 | -0.30, -0.11 | <0.001 |
| *Some college/associate's degree* | -0.28 | -0.37, -0.19 | <0.001 |
| *Bachelor's degree* | -0.39 | -0.48, -0.29 | <0.001 |
| *Postgraduate/professional degree* | -0.43 | -0.53, -0.33 | <0.001 |
| **Employment** |  |  |  |
| Working – paid or self-employed | ref | ref | ref |
| Not working – retired/disabled/other | 0.03 | 0.00, 0.07 | 0.079 |
| **Dental insurance** |  |  |  |
| Have dental insurance | ref | ref | ref |
| *Do not have dental insurance* | 0.16 | 0.12, 0.20 | <0.001 |

Ref = reference category; models control for age in years, race/ethnicity, highest education level achieved, annual household income, employment status, and dental insurance status; italicized row = significant at the $p < 0.05$ level; adjusted $R^2 = 0.09$ .

